# Large-scale accelerometry-derived physical activity and sedentary behaviour assessments in relation to health characteristics and outcomes among participants of the Lifelines Biobank study: rationale and design of the ActiveLIFE cohort study

**DOI:** 10.64898/2026.09.14.26362973

**Authors:** Milou Netten, Sabine Schootemeijer, Anna Sijtsma, Esmée A. Bakker, Thijs M.H. Eijsvogels

## Abstract

**Introduction:** Current guidelines on physical activity (PA) and sedentary behaviour (SB) are based on self-reported questionnaire data, which are prone to recall bias and measurement error. As such, they provide limited guidance on the intensity, frequency and duration of activities. To improve and accelerate research on the health benefits of a physically active lifestyle and inform future guidelines, large-scale accelerometry in population-based studies is needed. The ActiveLIFE cohort study aims to create a comprehensive, deeply-phenotyped dataset of thigh- and wrist-worn accelerometry in relation to health characteristics and outcome data that can be used to accelerate lifestyle and prevention research.

**Methods and analysis:** ActiveLIFE is an add-on study to the 4^th^ assessment wave of the Lifelines Biobank study, aiming to collect ∼60,000 accelerometry measurements. Lifelines participants aged ≥8 years old will be invited to wear a triaxial thigh-worn accelerometer (Axivity AX3, settings: 100 Hz, 8g) for 8 consecutive days. Morover, to evaluate how wear location impacts effect estimates, a subsample of at least 10% will be asked to wear both a thigh- and wrist-worn accelerometer. Raw accelerometry data will be processed into PA metrics, including steps per day, time spent in moderate-to-vigoruous PA, light-intensity PA, SB, and sleep and enriched with data on demographics, body composition, health status, medication use, lifestyle characteristics, biomarkers, cognitive function, cardiopulmonary performance, genetics, healthcare consumption and health outcome data. ActiveLIFE will allow to further refine, extend and personalize future guidelines on PA and SB for optimal PA prescription.

**Ethics & dissemination:** Ethics approval for the Lifelines Biobank, including the ActiveLIFE add-on study, was obtained from the Medical Ethics Committee of the University Medical Centre Groningen, the Netherlands (#2007/152). Informed consent was obtained from all participants. Study findings will be disseminated through peer-reviewed publications and conference presentations, and societal outreach approaches.

**What is already known:**

- Regular physical activity reduces the risk for mortality and major non-communicable diseases.
- Current physical activity guidelines are based on self-reported questionnaires, which are prone to recall bias, provide inaccurate estimates of activity intensity and volume, and do not capture all activity types.

**What this study adds:**

- ActiveLIFE will establish one of the largest deeply-phenotyped cohorts combining 24-hour thigh-worn accelerometry with extensive health, lifestyle, genetic, and clinical data.
- A subsample with simultaneous thigh- and wrist-worn accelerometry will enable direct comparison of wear locations and their impact on physical activity estimates and associations with health outcomes.

**How this study might affect research, practice, or policy:**

- Findings of the ActiveLIFE study will increase understanding of the association between physical activity and health outcomes and contribute to the refinement and optimalization of physical activity recommendations.
- The dual-placement (thigh- and wrist) allows to guide translation of physical activity recommenations to consumer wearables.

## Background and rationale

Regular physical activity (PA) reduces the risk for mortality, as well as major non-communicable diseases, including cardiovascular, neurological, and respiratory diseases [1]. Therefore, the current World Health Organization (WHO) guideline on PA and sedentary behaviour (SB) recommends adults to perform at least 150 minutes of moderate-to vigorous-intensity PA (MVPA) per week, to conduct strength training twice per week, and to reduce sedentary time [2]. The evidence driving these recommendations is mostly based on self-reported questionnaire data [2], which are prone to misreporting, and provide inaccurate estimates of activity intensity and duration [3, 4]. Furthermore, questionnaires cannot capture light-intensity PA (LIPA), vigorous intermittent lifestyle PA (VILPA; brief episodes of vigorous activity embedded within everyday activities), step-based parameters (including step count, cadence, and stepping patterns), or PA micropatterns (i.e., the timing, frequency, duration, and sequencing of activity and sedentary bouts), despite growing evidence that these metrics provide additional insights into health beyond total physical activity volume [5-7]. Accelerometers can overcome these limitations of self-report by continouously capturing PA and SB at a high resolution under real-world conditions [8], allowing accurate detection of postures and PA sub-types such as cycling [9, 10].

Therefore, the ActiveLIFE cohort study aims to create a large-scale comprehensive dataset of thigh- and wrist-worn accelerometry in relation to deeply phenotyped health and health outcome data. This data source can be used by researchers, clinicians and policy makers to increase our understanding of PA and SB and contribute to the refinement and optimalization of PA and SB recommendations.

## Methods

### Study population and setting

The ActiveLIFE study will be performed within the Lifelines Biobank cohort study and will leverage its existing infrastructure. Lifelines is a large, multi-generational, prospective cohort study that was initiated in 2006 and includes over 167,000 participants from the northern part of the Netherlands. An extensive description of the Lifelines study population, baseline data collection and follow-up procedures have been published previously [11, 12]. In brief, individuals aged 25-50 from the northern provinces of the Netherlands were invited by their general practitioner to participate in the Lifelines study between 2006 and 2013. On inclusion, the participants’ children and parents were also invited to participate, ensuring a three-generational cohort. Participants in Lifelines receive follow-up questionnaires every 1.5-2.5 years and visit the research centre every 5 years for follow-up data collection. As part of a lifelong cohort study, Lifelines is designed to track participants indefinitely.

ActiveLIFE will be embedded in the 4^th^ assessment wave of Lifelines. We aim to conduct 60,000 accelerometery measurements between July 2025 and December 2028. Accelerometery will be embedded in the standard measurements in the 4^th^ wave of Lifelines. All Lifelines participants aged 8 years and older will undergo the accelerometry measurement, except when they opt-out. We will document the reasons for opting out (**Table 1**). Participants will be excluded if they have an allergy to one or more of the materials used for the measurement (i.e. the accelerometer, wristband or adhesive film). To stimulate participation, participants will receive information about the accelerometry measurement prior to their visit to the Lifelines research centre, including an explanatory video (https://youtu.be/OqYRWyvl58U) describing the importance of wearing the accelerometer and explaining the measurement procedure.

**Table 1.** Reasons for opting out of accelerometer measurement participation.

| Reason category | Description |
| --- | --- |
| Participant preference | Participant does not wish to wear the accelerometer |
| Device-related | Device is perceived as uncomfortable to wear |
| Perceived usefulness | Measurement is not considered useful by the participant |
| Skin-related concerns | Skin sensitivity or concerns about skin irritation |
| Scheduling reasons | Measurement timing is inconvenient (e.g. participant will be on vacation during the measurement period) |
| Other | Other reason(s), specified by the participant |

### Accelerometry data collection

Twenty-four-hour PA patterns will be assessed objectively using the Axivity AX3 triaxial accelerometer (Open Lab, Newcasle University, UK). The Axivity AX3 accelerometer is validated for wearing on both the wrist and thigh [13, 14] and has been used in several other large population-based PA studies [15-17] (**Table 2**). The devices are waterproof but must be removed in the sauna, while deep-sea diving, and when undergoing an MRI scan. The Axivity AX3 will be set to measure at a dynamic range of ± 8 g and record data at 100 Hz. The AX3 OmGui software (version 1.0.0.37; Open Movement, Newcastle University, United Kingdom) will be used to configure the sensors and download the accelerometer data. Accelerometers will be configured up to one week in advance and programmed to start recording at 23:59 on the day of the participant’s visit. The devices will be configured individually, as each accelerometer will be assigned a unique code that will be linked to the participant during their visit. After the devices are returned, raw accelerometer data will be downloaded from the devices batch-wise and stored on the Lifelines servers.

**Table 2.**
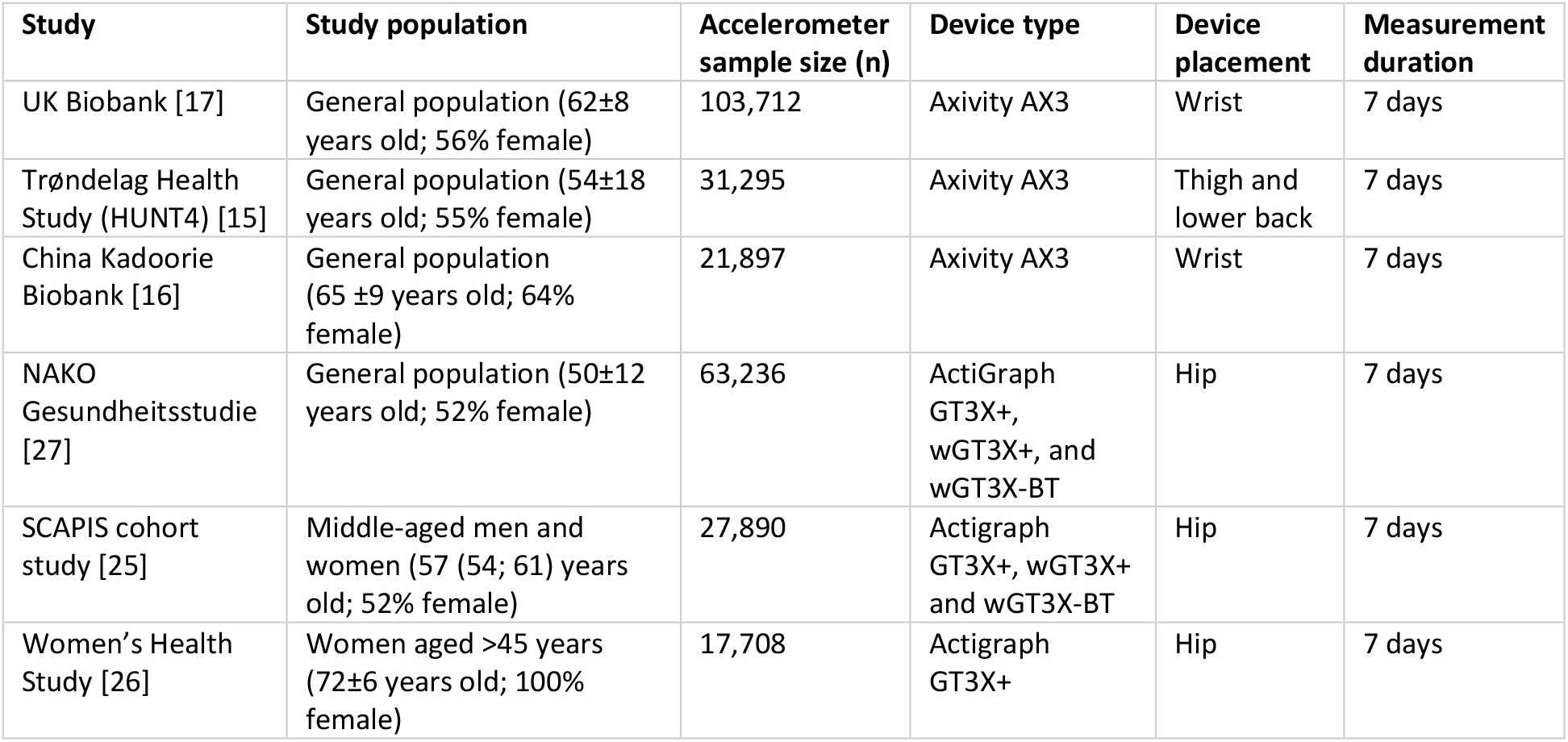
Overview of large population-based cohort studies (>10.000 participants) including accelerometer measurements.

Default accelerometer placement will be on the right thigh. Concurrent collection of thigh-worn and wrist-worn accelerometery will be conducted in a subsample of at least 10%. All partcipants aged ≥ 12 years will be eligible for dual measurements, as the wrist-worn device is not suitable for participants aged <12 years due to its size. Research nurses will attach the accelerometers during the participants’ first visit to the research centre. The thigh-worn accelerometers will be attached using moisture permeable adhesive film (Opsite Flexifix; Smith & Nephew, Watford, United Kingdom) on the right thigh, 10 cm above the kneecap. First, a 8*10cm piece of adhesive film will be placed on the thigh, followed by the device and another 8*10cm piece of adhesive film (**Figure 1**). The wrist-worn accelerometers will be attached using an accompanying wrist-band (Axivity Ltd., Newcastle, UK) on the non-dominant wrist. Participants will receive an extra set of adhesive film to reattach the device themselves in case it starts to loosen. Participants will wear the accelerometer continuously for 8 days, including when showering or swimming (**Figure 2**). We chose a measurement period of 8 days as previous studies showed that measurement periods between 7 and 10 days are necessary to accurately capture MVPA and sedentary time [18, 19]. Participants will return the accelerometer during their second visit to the research centre or by mail if the second visit occurred within <7 days or > 21 days following the first study visit. Participants who have not returned the device after 21 days will be contacted by telephone and asked to return the device by mail.

**Figure 1.**
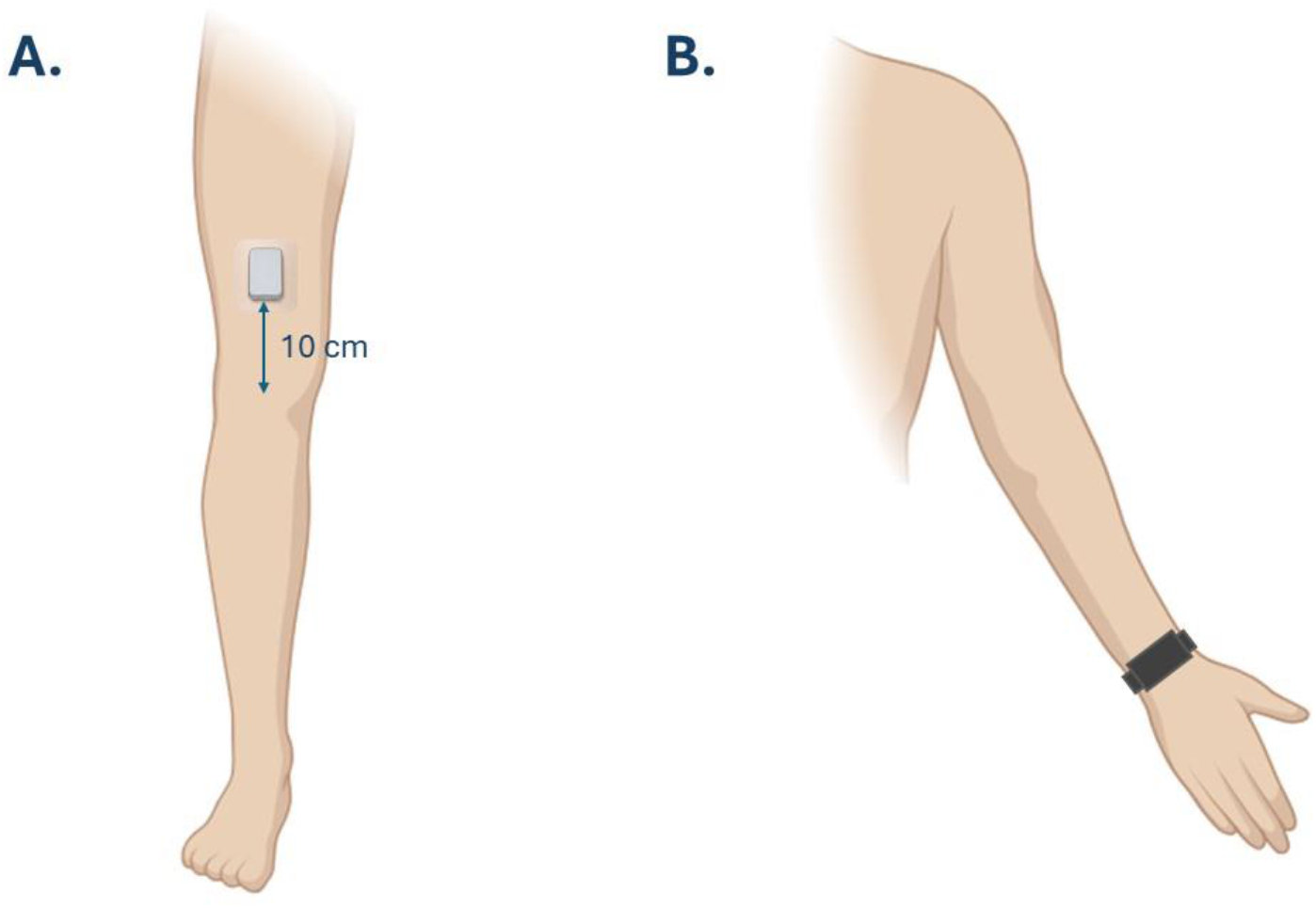
Accelerometer placement on the right thigh (A) and on the non-dominant wrist (B).

**Figure 2.**
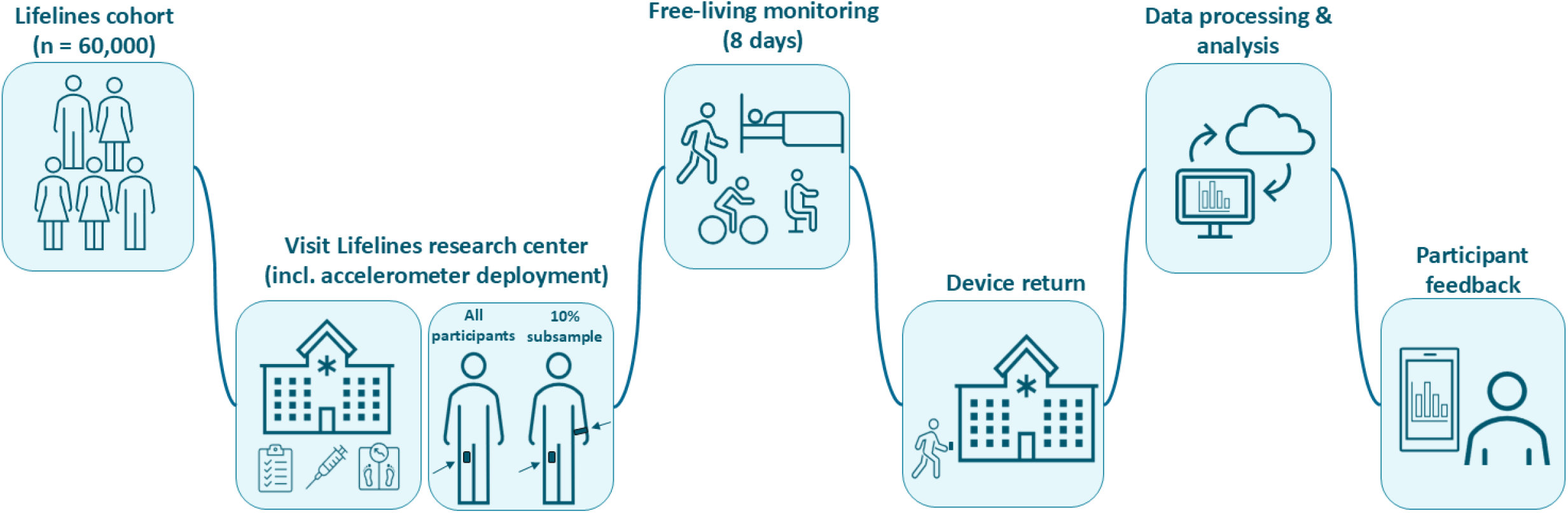
Overview of the accelerometer measurement protocol in the Lifelines Cohort (n expected = 60,000).

After the accelerometer is returned and the data is extracted and processed, participants will receive short and simple feedback on their PA volume (mean MVPA (min/day), step count (steps/day) and sedentary time (hours/day)).

### Post-processing raw accelerometer data

Raw accelerometer data will be processed with the validated ActiPASS algorithm (Version 2025.04) for the thigh-worn accelerometer data, and the UK Biobank Accelerometer algorithm (Version 7.5.0) for the wrist-worn accelerometer data [13, 20-23]. Days will be classified as non-valid when they have <20h of wear time, 0 minutes of walking, or 0 minutes of sleep [24]. Participants with no valid days will be removed from the dataset. PA metrics will include steps per day, time spent in MVPA, LIPA, SB, and sleep. The full list of available accelerometery variables can be found on the Lifelines Wiki (https://edu.nl/7ngbf). Raw accelerometer data will be stored and made available to other researchers, to facilitate the use of different or updated algorithms in the future.

To assess the validity of the selected algorithms for classifying PA types and posture in our study population, we will conduct an activity-recognition validation study. Further details on the rationale, design and outcomes of this sub-study are described elsewhere (https://edu.nl/wk6d9).

### Data integration

Accelerometer data collected during the 4^th^ assessment wave of Lifelines will be linked to the extensive longitudinal information already available from the 1^st^ to 3^rd^ assessment waves (2007–2023). These data include body composition, health status, medication use, lifestyle characteristics (e.g. diet, living environment), blood and urine biomarkers, cognitive function, cardiopulmonary performance, and genetic information (**Figure 3**). A complete overview of available variables can be found in the Lifelines data catalogue (https://edu.nl/877rw). The accelerometer assessment in the 4^th^ wave will be accompanied by the routine Lifelines examinations and questionnaires, allowing concurrent characterization of participants with respect to their health characteristics. Prospective health outcomes will be obtained through ongoing Lifelines follow-up measurements and linkage with external registries. Health outcomes, including cause-specific mortality, healthcare utilization and expenditures, will be derived through linkage with Statistics Netherlands (https://edu.nl/x4kvc).

**Figure 3.**
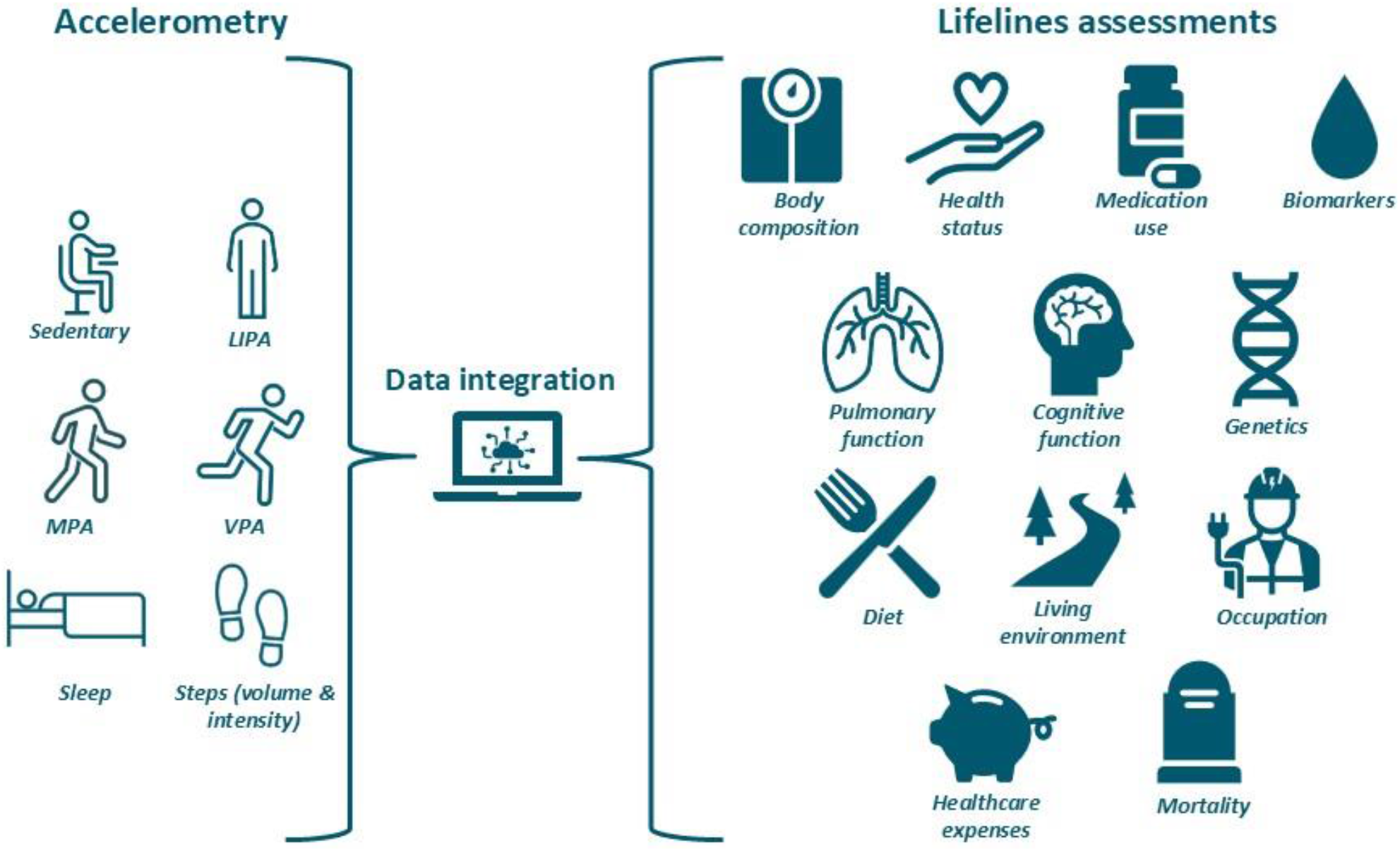
Integration of data collected in ActiveLIFE and Lifelines. LIPA = light-intensity physical activity, MPA = moderate-intensity physical activity, VPA = vigorous intensity physical activity.

### Data management and access

The project will carefully follow all GDPR requirements to protect generated data. All data will be collected and stored according to the current scientific standards and Good Clinical Practice guidelines. Raw and processed accelerometer files will be stored on the Lifelines servers and will be linked to participants using pseudonymized identifiers. Directly identifiable information will not be made available to researchers.

Processed and raw accelerometer data, data on health and well-being, and corresponding metadata will be made available to other researchers using the Lifelines Catalogue infrastructure. External researchers will be able to request access to the data through the established Lifelines application procedure (https://edu.nl/f9arj). Approved datasets will be provided within a secure digital research environment (e.g. anDREa (Analytics Environment for Data Research and Analysis; https://edu.nl/nde76)), where researchers will be able to access and analyse data. Future analysis plans using the ActiveLIFE data will be published on the Open Science Framework (https://edu.nl/xvdtx). Following this work flow, the ActiveLIFE cohort fully complies with the Open Science principles including FAIR data management, reproducibility, and inclusive, collaborative practices.

### Ethics & dissemination

Ethical approval for the Lifelines Biobank, including the ActiveLIFE add-on study, was obtained from the Medical Ethics Committee of the University Medical Centre Groningen, the Netherlands (#2007/152). Lifelines is conducted according to the principles of the Declaration of Helsinki and informed consent was obtained from all participants. The results of the ActiveLIFE study will be disseminated through peer-reviewed articles, presentations at (inter)national conferences and societal outreach approaches.

## Discussion

In the ActiveLIFE cohort study, we aim to perform 60,000 thigh – and wrist-worn accelerometery measurements in a population-based cohort. Accelerometry data will be combined with extensive assessments, including demographics, body composition, health status, medication use, lifestyle characteristics (e.g. diet, living environment), biomarkers, cognitive function, cardiopulmonary and physical performance, genetic information, as well as healthcare consumption and health outcomes. This rich datasource will allow researchers, clinicians and policy makers to establish dose–response relationships between PA and SB metrics, and health outcomes. We will be able to examine the health effects across the full 24-hour spectrum, modelling the potential health effects of various lifestyle changes (i.e. replacing SB with LIPA or MVPA), and developing personalized PA and SB recommendations.

The ActiveLIFE cohort will be a valuable addition to the expanding landscape of population-based studies with wearable data (**Table 2**). A unique element of ActiveLIFE is that objective measures of 24-hour movement behaviour will be primarily derived from thigh-worn accelerometers, versus wrist-worn [16, 17] and hip-worn [25-27] in other cohorts. The reason for this is that thigh-worn accelerometry is most reliable at capturing common activities such as SB and cycling compared to wrist- or hip-worn [9, 10]. Another innovative aspect is that, a subset of at least 10% of participants will undergo dual-sensor placement (thigh and wrist). The dual-placement will allow to directly compare device locations, improve harmonisation with studies that rely solely on wrist-worn accelerometery and facilitate translation to consumer wearables, such as smartwatches.

The ActiveLIFE study has several strengths. First, ActiveLIFE will yield a comprehensive and deeply phenotyped dataset of thigh-worn accelerometery combined with extensive health assessments. PA behaviour will be collected at high-resolution in each participant, with continuous monitoring over eight days at a sampling frequency of 100 Hz. Second, the large sample size of ±60,000 measurements will allow to conduct subgroup analyses with sufficient statistical power. Third, ActiveLIFE will benefit from the existing Lifelines infrastructure that was developed over the past 15 years, making the project highly feasible[11, 12].

We also anticipate several challenges. First, we are aiming for 60,000 accelerometer measurements among the expected 80,000 participants of the 4^th^ assessment wave of the Lifelines Biobank study. This requires a response rate of at least 75%. Response rates in other large-scale population based cohort studies vary greatly, between 45% and 81% [15-17]. To ensure a high response rate for the ActiveLIFE study, accelerometery will be part of the core measurements of the 4^th^ Lifelines assessment wave, with an opt-out approach. As of September 2026 (n= 16,321), we have a response rate of 85%, so we expect to reach our target of 60,000 measurements. Second, the use of thigh-worn accelerometery may reduce translation of findings to wrist-worn consumer wearables like bracelets and smartwatches. However, we deliberately opted for thigh-worn accelerometery as this wear-location yields the most reliable estimates for SB (i.e. sitting, lying), standing, walking and cycling [9, 10]. We will simultaneously collect thigh-worn and wrist-worn accelerometery in a sub-sample of our study population, which will allow to evaluate the impact of wear location and to translate project findings to wrist-worn consumer- and research wearables. Third, the Lifelines study participants were recruited exclusively from the northern Netherlands which may limit representativeness for the Dutch population. However, the sample closely reflects the demographics of the general population with respect to age (Lifelines: 41 years, general population: 43 years), sex (Lifelines percent women: 58.5%, general population percent women: 50.7%), education level (Lifelines percent highly educated: 28.8%, general population percent highly educated: 22,8%), and chronic disease prevalence (<3% difference between Lifelines and general population;[11, 28, 29]).

In conclusion, the ActiveLIFE study will create a large and deeply phenotyped accelerometery dataset. This dataset will be used to investigate the dose-response relationship between PA and SB characteristics, including novel PA metrics, with health outcomes. The ultimate goal of ActiveLIFE is to advance from generic-to personalized and disease-specific PA recommendations.

### Current status

Data collection for the ActiveLIFE study was iniatied on July 1^st^ 2025. As of September 2026, 16,321 accelerometry measurements have been performed. Updates on data collection progress will be posted regularly on the ActiveLIFE webpage (https://edu.nl/bge9c). We expect to finish data collection by December 2028.

## Data Availability

All data produced in the present study are available upon reasonable request to the authors

## References

1. Lee, I.M., et al., Effect of physical inactivity on major non-communicable diseases worldwide: an analysis of burden of disease and life expectancy. Lancet, 2012. 380(9838): p. 219–29.

2. Bull, F.C., et al., World Health Organization 2020 guidelines on physical activity and sedentary behaviour. Br J Sports Med, 2020. 54(24): p. 1451–1462.

3. Sember, V., et al., Validity and Reliability of International Physical Activity Questionnaires for Adults across EU Countries: Systematic Review and Meta Analysis. Int J Environ Res Public Health, 2020. 17(19).

4. Gill, J.M., et al., Potential impact of wearables on physical activity guidelines and interventions: opportunities and challenges. Br J Sports Med, 2023. 57(19): p. 1223–1225.

5. Stamatakis, E., et al., Association of wearable device-measured vigorous intermittent lifestyle physical activity with mortality. Nat Med, 2022. 28(12): p. 2521–2529.

6. Stamatakis, E., et al., Dose Response of Incidental Physical Activity Against Cardiovascular Events and Mortality. Circulation, 2025. 151(15): p. 1063–1075.

7. Stens, N.A., et al., Relationship of Daily Step Counts to All-Cause Mortality and Cardiovascular Events. J Am Coll Cardiol, 2023. 82(15): p. 1483–1494.

8. Migueles, J.H., et al., Accelerometer Data Collection and Processing Criteria to Assess Physical Activity and Other Outcomes: A Systematic Review and Practical Considerations. Sports Med, 2017. 47(9): p. 1821–1845.

9. Montoye, A.H.K., et al., Validation and Comparison of Accelerometers Worn on the Hip, Thigh, and Wrists for Measuring Physical Activity and Sedentary Behavior. AIMS Public Health, 2016. 3(2): p. 298–312.

10. Bach, K., et al., A Machine Learning Classifier for Detection of Physical Activity Types and Postures During Free-Living. Journal for the Measurement of Physical Behaviour, 2022. 5(1): p. 24–31.

11. Scholtens, S., et al., Cohort Profile: LifeLines, a three-generation cohort study and biobank. International Journal of Epidemiology, 2015. 44(4): p. 1172–1180.

12. Sijtsma, A., et al., Cohort Profile Update: Lifelines, a three-generation cohort study and biobank. International Journal of Epidemiology, 2022. 51(5): p. E295–E302.

13. Crowley, P., et al., Comparison of physical behavior estimates from three different thigh-worn accelerometers brands: a proof-of-concept for the Prospective Physical Activity, Sitting, and Sleep consortium (ProPASS). Int J Behav Nutr Phys Act, 2019. 16(1): p. 65.

14. Brocklebank, L., et al., Comparing Two Wrist-Worn Accelerometers (Axivity AX3 and Matrix 003) for Measuring Movement Behaviors in British and Chinese Older Adults. Journal for the Measurement of Physical Behaviour, 2025. 8(1).

15. Asvold, B.O., et al., Cohort Profile Update: The HUNT Study, Norway. Int J Epidemiol, 2023. 52(1): p. e80–e91.

16. Chen, Y., et al., Device-measured movement behaviours in over 20,000 China Kadoorie Biobank participants. Int J Behav Nutr Phys Act, 2023. 20(1): p. 138.

17. Doherty, A., et al., Large Scale Population Assessment of Physical Activity Using Wrist Worn Accelerometers: The UK Biobank Study. PLoS One, 2017. 12(2): p. e0169649.

18. Aadland, E. and E. Ylvisaker, Reliability of Objectively Measured Sedentary Time and Physical Activity in Adults. PLoS One, 2015. 10(7): p. e0133296.

19. Hilden, P., et al., How many days are needed? Measurement reliability of wearable device data to assess physical activity. PLoS One, 2023. 18(2): p. e0282162.

20. Doherty, A.C. S.; Yuan, H.; Walmsley, R., accelerometer: A Python Toolkit for Extracting Physical Activity and Behavior Metrics from Wearable Sensor Data. 2020.

21. Willetts, M., et al., Statistical machine learning of sleep and physical activity phenotypes from sensor data in 96,220 UK Biobank participants. Sci Rep, 2018. 8(1): p. 7961.

22. Hettiarachchi, P.J. P., ActiPASS. 2025.

23. Stemland, I., et al., Validity of the Acti4 method for detection of physical activity types in free-living settings: comparison with video analysis. Ergonomics, 2015. 58(6): p. 953–65.

24. Blodgett, J.M., et al., Device-measured physical activity and cardiometabolic health: the Prospective Physical Activity, Sitting, and Sleep (ProPASS) consortium. Eur Heart J, 2024. 45(6): p. 458–471.

25. Ekblom-Bak, E., et al., Accelerometer derived physical activity patterns in 27.890 middle-aged adults: The SCAPIS cohort study. Scand J Med Sci Sports, 2022. 32(5): p. 866–880.

26. Lee, I.M., et al., Using Devices to Assess Physical Activity and Sedentary Behavior in a Large Cohort Study, the Women’s Health Study. J Meas Phys Behav, 2018. 1(2): p. 60–69.

27. Peters, A., et al., Framework and baseline examination of the German National Cohort (NAKO). Eur J Epidemiol, 2022. 37(10): p. 1107–1124.

28. (CBS), C.B.v.d.S., Bevolking op 1 januari en gemiddeld; geslacht, leeftijd en regio. 2025.

29. Klijs, B., et al., Representativeness of the LifeLines Cohort Study. PLoS One, 2015. 10(9): p. e0137203.

